# Large-scale analysis demonstrates the influence of CYP2C19 genotype on specific SSRI side effects

**DOI:** 10.1101/2024.12.20.24319269

**Authors:** Chris Eijsbouts, Yunxuan Jiang, James Ashenhurst, Julie M. Granka, 23andMe Research Team, Steven Pitts, Adam Auton, Noura S. Abul-Husn, Alison Chubb, R. Ryanne Wu

**Affiliations:** 23andMe, Inc., 223 N. Mathilda Ave., Sunnyvale, CA 94086

**Author notes:** Corresponding authors: Chris Eijsbouts, PhD, R. Ryanne Wu, MD.

## Abstract

The majority of individuals receiving treatment for major depressive disorder (MDD) do not achieve remission from the first medication they try, and over 80% subsequently discontinue pharmacotherapy or switch to a different medication. SSRI discontinuation due to side effects is common.

We evaluated the effect of CYP2C19 genotype on SSRI response using self-reported data from 114,627 direct-to-consumer genetics research participants who were prescribed an SSRI primarily metabolized by CYP2C19 (citalopram, escitalopram, or sertraline).

Among participants taking citalopram or escitalopram, slower metabolizers experienced side effects significantly more often than faster metabolizers (OR=1.04 per grade, from 0 for poor metabolizers to 5 for ultrarapid metabolizers, 95%CI=[1.02-1.06] and OR=1.05 per grade, 95%CI=[1.02-1.07]) and were more likely to discontinue treatment due to side effects (OR=1.05, 95%CI=[1.03-1.08], e.g. 29.7% of poor vs. 21.6% of ultrarapid metabolizers, and OR=1.07, 95%CI=[1.04-1.11], e.g. 25.7% vs. 20.2%). Slower metabolizers taking escitalopram were more likely to suffer from sleep problems and sexual problems than faster metabolizers. Slower metabolizers taking sertraline reported tremor more often than faster metabolizers.

Overall, we find substantial differences in side effect risk between individuals with different CYP2C19 genotypes in a large sample, supporting the notion that individuals seeking treatment for MDD may benefit from preemptive pharmacogenetic testing and genotype-guided dosing recommendations to minimize side effects and reduce discontinuations.

## Introduction

Alongside an increased prevalence of anxiety and major depressive disorder (MDD) in the past decade,^1,2^ rates of antidepressant use have risen, particularly in recent years.^3,4^ Antidepressant use in the United States is now widespread: more than 1 in 10 adults and adolescents reported recent antidepressant use – a rate that climbs to more than 1 in 5 among women over forty.^5,6^ The majority (62%) of individuals starting antidepressant monotherapy are prescribed selective serotonin reuptake inhibitors (SSRIs).^7^ Remission rates following first-line SSRI treatment for MDD are low with less than half of patients responding, and only 36.8% achieving remission.^8^ Within a year of starting SSRI treatment, roughly 80% of patients switch treatments or discontinue pharmacological treatment altogether – only 20% persist for a year with their first SSRI.^7^ After up to four treatment re-evaluations (i.e. opportunities to switch treatments), closer to 70-90% of patients receiving treatment may ultimately achieve remission,^9^ typically requiring a lengthy trial-and-error process. Among patients treated with SSRIs, side effects are a frequent reason to switch treatments.^10^ In short, finding an appropriate antidepressant remains a challenge for a large portion of the US population.

Considering the effects of MDD on quality of life^11^ and its significant economic impact, with costs in the United States exceeding 380 billion USD (2023) annually,^7^ novel approaches to help patients and physicians find appropriate treatments more quickly are needed. Genetics-guided treatment using pharmacogenetic testing is one area of potential promise for antidepressant prescribing. Most SSRIs, for example, are predominantly metabolized by the cytochrome P450 (CYP) liver enzyme CYP2C19, and variants in the CYP2C19 gene can significantly alter the enzyme’s function. Knowing if a patient has these variants and choosing treatments and dosing based on predicted CYP2C19 metabolizer status has shown promise in improving response rates, remission rates,^12^ and time to titration.^13^ While randomized clinical trials typically find a consistent, positive effect of pharmacogenetics-guided MDD treatment on the odds of remission, obtaining sufficient power to detect statistically significant effects remains an issue, particularly over longer treatment periods, where data availability is more limited.^14,15^

Previous studies have been underpowered to detect associations between CYP2C19 metabolizer status and specific SSRI-related side effects. In large genotyped or sequenced cohorts, such as UK Biobank, data on SSRI-related side effects and efficacy remain sparse. Recent studies have typically pooled side effects to maximize statistical power, oftentimes still not detecting effects of CYP2C19 genotype (or the metabolizer status predicted from it) on drug response. Wong et al.^16^ observed no associations between CYP2C19 status and escitalopram discontinuation, and no associations with citalopram that survived multiple testing correction. Other recent work does not appear powered to detect associations between CYP2C19 metabolizer status and citalopram or escitalopram side effects (when pooled), or with the efficacy of any single SSRI.^17^

Here, we present the largest analysis of the effect of CYP2C19 genotype on SSRI drug response to date, using self-reported data from 114,627 consumer genetics research participants (23andMe, Inc., Sunnyvale, CA) who were prescribed an SSRI primarily metabolized by CYP2C19.

## Methods

### Survey data on depression treatments

Research participants were customers of 23andMe, Inc., a consumer genetics and research company in Sunnyvale, CA, who provided informed consent and volunteered to participate in the research online, under a protocol approved by the external Association for Accreditation of Human Research Protection Programs (AAHRPP)-accredited IRB, Ethical & Independent (E&I) Review Services. As of 2022, E&I Review Services is part of Salus IRB (https://www.versiticlinicaltrials.org/salusirb). Treatment response data were collected between March 2022 and July 2024 from a survey about depression treatments. Research participants who previously reported experiencing depression or who indicated having received a clinical diagnosis of depression (via 23andMe’s current Health Profile survey, Health Update survey, or previous 23andMe mental health surveys) were eligible to complete the depression treatment survey, and were asked to confirm that they had received a diagnosis of depression. To be included in this study, research participants needed to report using or having used escitalopram, citalopram and/or sertraline, as identified using either their generic name or brand name. All participants who had ever taken these treatments were asked about (1) medication efficacy, (2) side effect(s), if experienced, and (3) reason(s) for medication discontinuation if applicable (Supplementary Note).

### CYP2C19 genotyping, predicted CYP2C19 metabolizer status assignment, and genetic ancestry classification

Sample collection, genome-wide genotyping, and CYP2C19 genotyping methods have been described previously.^18^ CYP2C19 metabolizer status (poor, intermediate, normal, rapid, and ultrarapid) was assigned based on the presence or absence of the three most common CYP2C19 alleles (*2, *3, and *17) and followed standard CYP2C19 genotype-metabolizer status mapping, implemented via an in-house method.^19,20^ As a cross check, we verified that the frequencies of each predicted metabolizer type closely matched (to within 1 percentage point, in all cases) those produced by PharmCat version 2.8.3 in a set of 175,026 customers that overlapped with the study sample.^21^

Additionally, genome-wide genotype data were used to assign each individual to a genetic ancestry group via a previously described classification algorithm,^22,23^ enabling SSRI response analyses per ancestry group.

### Data analysis

To test associations between drug response phenotypes and CYP2C19 metabolizer status, we graded each CYP2C19 metabolizer status based on its metabolic efficiency (on a five point scale, from 0 for ultrarapid metabolizers to 4 for poor metabolizers), and regressed the drug response phenotypes on these grades using logistic regression separately in each ancestry group. We then meta-analyzed results across ancestry groups. Odds ratios reported for these associations therefore reflect the change in drug response (e.g. an increased chance of discontinuing due to side effects) per one step change in metabolizer speed (e.g. a change from rapid to normal metabolizer). We chose this strategy to maximize statistical power, as it avoids the discard of data that would be required for a pairwise analysis (e.g., comparing only normal metabolizers to only rapid metabolizers).

While side effects and discontinuation were binary, symptom improvement was reported on a 5-point scale ranging from “much worse” to “much improved” symptoms. We coded symptom improvement in two different ways, (1) a 5-point scale from -2 for “much worse” symptoms to +2 for “much improved” symptoms, and (2) a 3 point-scale from -1 for symptoms that were either much or a little worse to +1 for symptoms that were either a little or much improved.

We carried out association testing using R version 4.2.1, separately for each genetic ancestry group, adjusting for differences in age, genetic sex, weight, and the first five principal components of genetic ancestry in that group. Cross-ancestry test statistics were obtained through fixed-effect inverse variance weighted meta-analysis, implemented in the meta R package, version 4.8-2.^24^ During meta-analysis, we excluded estimates derived from models that failed to converge, or where we observed (quasi-)complete separation. These issues only affected estimates in the Middle Eastern and South Asian ancestry groups, likely due to their limited sample size. When analyzing side effects, we pooled data from past and current users of each SSRI. Analyses of discontinuation were, by definition, limited to past SSRI users.

## Results

### Participant demographics

114,627 23andMe research participants completed the depression treatment survey. Most were of European ancestry (80.9%) and most were female (82.9%). Roughly a fifth (20.3%) of participants had been previously hospitalized for symptoms related to depression, with the highest hospitalization rates found in the African American genetic ancestry group (e.g., 34.9% among African American citalopram users). Most had taken at least two antidepressants (63.3%, IQR 1-4, median 2) at the time of the survey. In line with previous literature,^18^ we observed pronounced cross-ancestry variation in metabolizer type frequencies (e.g., 17.8% or 98/551 of East Asian-ancestry escitalopram users were poor CYP2C19 metabolizers, versus 2.3% or 1,060/45,795 of European-ancestry escitalopram users). Demographics of respondents are detailed in **Table 1**.

**Table 1:** Research participant demographics, stratified by SSRI and genetic ancestry group. “% Female” refers to genetic sex. “Antidep. taken” refers to the number of different antidepressants participants have taken, in the present and in the past, to treat depression (see Supplementary Note, only recognized medications were counted, “other” treatments were excluded). The last five columns show how common a particular metabolizer status is. PM: Poor metabolizer; IM: Intermediate Metabolizer; NM: Normal Metabolized; RM: Rapid Metabolizer; UM: Ultrarapid Metabolizer. In order to respect 23andMe research participant privacy, we do not report metabolizer frequency percentages on rows where some of these would correspond to groups of five or fewer individuals.

| SSRI | Genetic ancestry | N | Female (%) | Age (mean) | Age (SD) | Weight (kg, mean) | Weight (SD) | Hospitalized (%) | Antidep. taken (median) | PM (%) | IM (%) | NM (%) | RM (%) | UM (%) |
| --- | --- | --- | --- | --- | --- | --- | --- | --- | --- | --- | --- | --- | --- | --- |
| Any | All | 114627 | 82.9 | 45.1 | 15.8 | 82.9 | 24.9 | 20.3 | 2 | 2.4 | 25.6 | 41.2 | 26.5 | 4.3 |
| Escitalopram | All | 56551 | 83.3 | 44.0 | 15.2 | 82.7 | 24.6 | 22.3 | 3 | 2.4 | 25.6 | 41.0 | 26.6 | 4.4 |
| Citalopram | All | 31625 | 84.3 | 47.4 | 15.1 | 83.9 | 24.7 | 23.9 | 3 | 2.3 | 25.8 | 41.0 | 26.5 | 4.4 |
| Sertraline | All | 67046 | 84.3 | 44.2 | 15.6 | 82.8 | 25.2 | 22.8 | 3 | 2.5 | 25.7 | 41.2 | 26.4 | 4.3 |
| Escitalopram | African American | 1500 | 85.5 | 39.7 | 13.3 | 87.2 | 25.1 | 31.1 | 2 | 3.1 | 28.8 | 38.3 | 24.2 | 5.7 |
| Escitalopram | East Asian | 551 | 81.1 | 35.2 | 11.3 | 65.3 | 17.1 | 22.9 | 2 |  |  |  |  |  |
| Escitalopram | European | 45795 | 83.4 | 45.2 | 15.4 | 83.3 | 24.8 | 21.4 | 3 | 2.3 | 25.2 | 40.2 | 27.6 | 4.7 |
| Escitalopram | Latine | 6456 | 83.5 | 38.8 | 13.0 | 81.1 | 23.6 | 26.9 | 3 | 1.6 | 23.8 | 48.3 | 23.5 | 2.7 |
| Escitalopram | Middle Eastern | 132 | 66.7 | 38.4 | 13.8 | 74.3 | 19.4 | 17.6 | 2 |  |  |  |  |  |
| Escitalopram | Other | 1984 | 81.3 | 40.1 | 14.6 | 76.6 | 21.6 | 23.0 | 3 | 2.9 | 31.1 | 40.2 | 22.0 | 3.8 |
| Escitalopram | South Asian | 133 | 72.9 | 36.7 | 10.5 | 73.2 | 22.4 | 18.0 | 2 |  |  |  |  |  |
| Citalopram | African American | 686 | 88.3 | 42.8 | 12.9 | 89.7 | 24.2 | 34.9 | 3 | 2.9 | 28.1 | 37.5 | 25.2 | 6.3 |
| Citalopram | East Asian | 191 | 84.3 | 40.3 | 12.7 | 67.1 | 15.4 | 24.1 | 3 |  |  |  |  |  |
| Citalopram | European | 26613 | 84.3 | 48.2 | 15.1 | 84.3 | 24.8 | 22.9 | 4 | 2.3 | 25.5 | 40.4 | 27.2 | 4.6 |
| Citalopram | Latine | 3116 | 84.5 | 42.0 | 13.4 | 82.6 | 25.4 | 30.6 | 3 | 1.6 | 23.4 | 47.7 | 24.7 | 2.5 |
| Citalopram | Middle Eastern | 43 | 65.1 | 49.4 | 17.7 | 78.5 | 20.6 | 9.5 | 2 |  |  |  |  |  |
| Citalopram | Other | 928 | 80.9 | 44.9 | 15.0 | 79.0 | 21.1 | 22.9 | 3 | 2.4 | 33.5 | 40.6 | 20.0 | 3.4 |
| Citalopram | South Asian | 48 | 66.7 | 38.3 | 11.8 | 69.8 | 15.9 | 22.9 | 3 |  |  |  |  |  |
| Sertraline | African American | 1924 | 87.1 | 39.9 | 13.7 | 87.2 | 24.6 | 31.9 | 2 | 2.9 | 30.7 | 36.4 | 25.2 | 4.8 |
| Sertraline | East Asian | 650 | 83.2 | 36.1 | 12.4 | 65.2 | 15.8 | 24.7 | 2 |  |  |  |  |  |
| Sertraline | European | 54054 | 84.4 | 45.4 | 15.8 | 83.2 | 25.3 | 21.9 | 3 | 2.3 | 25.3 | 40.3 | 27.4 | 4.7 |
| Sertraline | Latine | 7860 | 84.6 | 38.8 | 13.1 | 81.9 | 25.7 | 27.1 | 2 | 1.9 | 23.6 | 48.4 | 23.6 | 2.6 |
| Sertraline | Middle Eastern | 158 | 64.6 | 39.0 | 14.2 | 74.1 | 18.1 | 15.3 | 2 |  |  |  |  |  |
| Sertraline | Other | 2268 | 82.0 | 40.9 | 15.2 | 77.1 | 21.7 | 23.2 | 2 | 3.9 | 30.2 | 42.1 | 20.7 | 3.0 |
| Sertraline | South Asian | 132 | 72.7 | 35.4 | 10.3 | 72.9 | 21.5 | 22.7 | 2 |  |  |  |  |  |

### SSRI side effects and discontinuation rates are similar across ancestry and sex

Around half of all participants who had ever used citalopram, escitalopram or sertraline reported one or more side effects (37.8%, 42.5% and 45.7%, respectively). Most side effects, such as sexual problems or sleep problems, were reported by around a fifth of participants, although rates of tremor were substantially lower (e.g., 3.9% among participants using sertraline, **Fig. 1, Supplementary Table 1**). The rate of experiencing any particular side effect was similar across all genetic ancestry groups for the three SSRIs evaluated. Participants who had taken citalopram, however, reported discontinuation due to a lack of efficacy notably more often (38.9%) than participants who were on either escitalopram (31.9%) or sertraline (29.6%).

**Figure 1:**
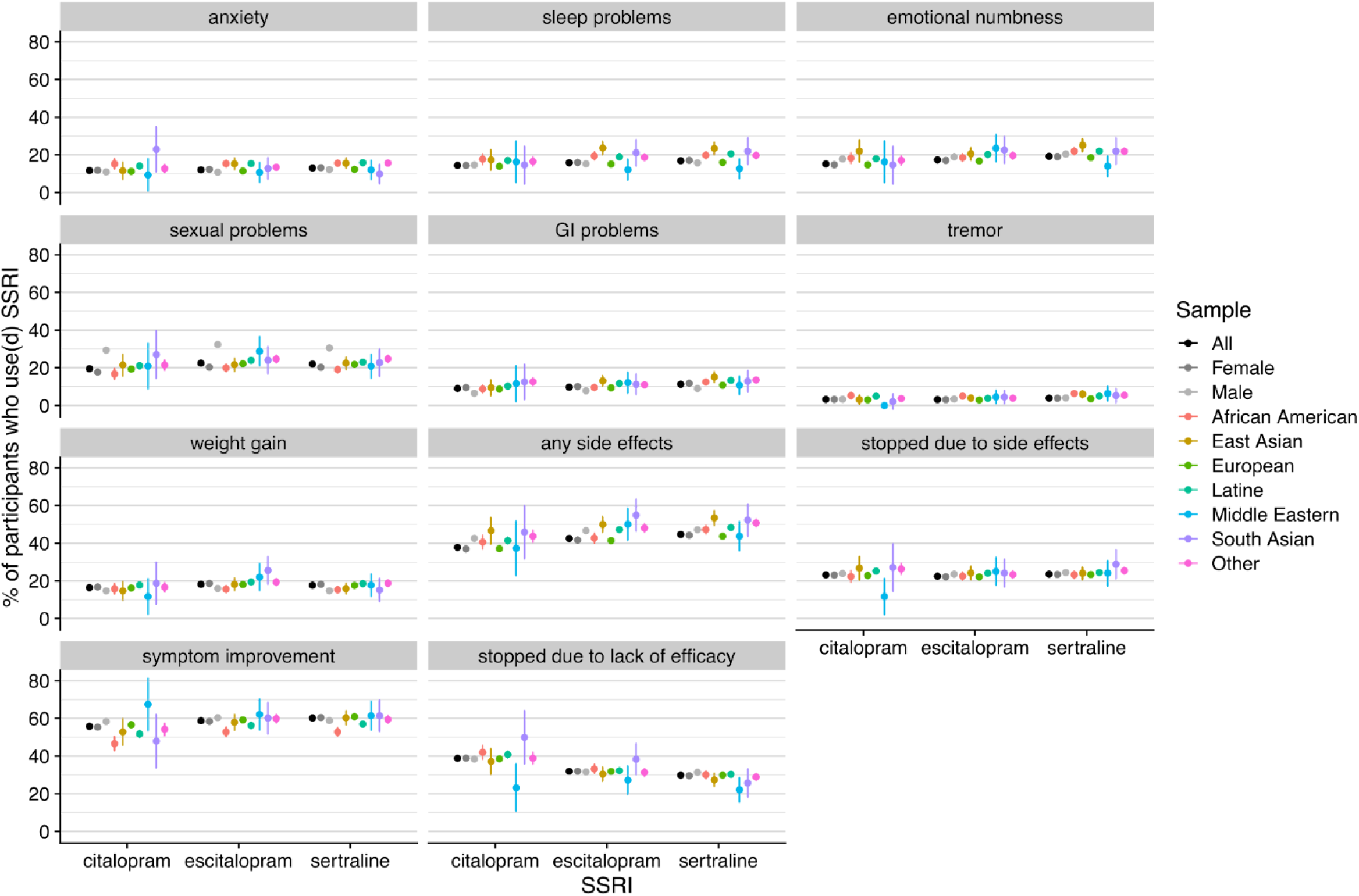
Prevalence of side effects and reasons for discontinuation of SSRI treatment. Shown per SSRI, stratified by sex and genetic ancestry group. “any side effects” refers to one or more of the listed side effects. “stopped due to side effects” and “stopped due to lack of efficacy” refer to reasons provided for treatment discontinuation. Vertical bars mark 95% confidence intervals. GI problems: gastrointestinal problems.

While side effects reporting was generally consistent between sexes, we observed a considerable difference with respect to sexual side effects: 32.3% of men who used or had used escitalopram reported sexual problems as a side effect versus 20.3% of women (OR = 0.48 for women, 95% CI [0.45-0.50] after adjusting for age, weight). Differences in sexual side effects between men and women were also observed with citalopram (29.3% vs 17.7%, OR=0.45, 95%CI [0.41-0.48]) and sertraline users (30.6% vs 20.3%, OR=0.50 95% CI [0.48-0.53]), which is consistent with previous reports.^25^

### CYP2C19 metabolizer status predicts individual SSRI side effects

Slower CYP2C19 metabolizers taking citalopram or escitalopram reported experiencing one or more side effects significantly more often than faster metabolizers after adjusting for age, sex, weight, and principal components of genetic background in the cross-ancestry meta-analysis (OR=1.04 per grade, 95%CI=[1.02-1.06] and OR=1.05 per grade, 95%CI=[1.02-1.07]) (**Fig. 2, Supplementary Table 2**). In general, slower CYP2C19 metabolizers tended to have an increased probability of experiencing any given individual side effect across all three SSRIs. After multiple-testing correction, among participants taking escitalopram, slower metabolizers were significantly more likely to suffer from sleep problems (OR=1.05, CI=[1.01-1.07], e.g. 18.5% of poor vs. 14.4% of ultrarapid metabolizers) and sexual problems (OR=1.04, 95%CI=[1.02-1.07], e.g. 24.4% vs. 20.6%) than faster metabolizers. Similarly, slower metabolizers taking sertraline reported tremor significantly more often (OR=1.10, 95%CI=[1.05-1.15], e.g. 6.3% vs. 3.1%) than faster metabolizers.

**Figure 2:**
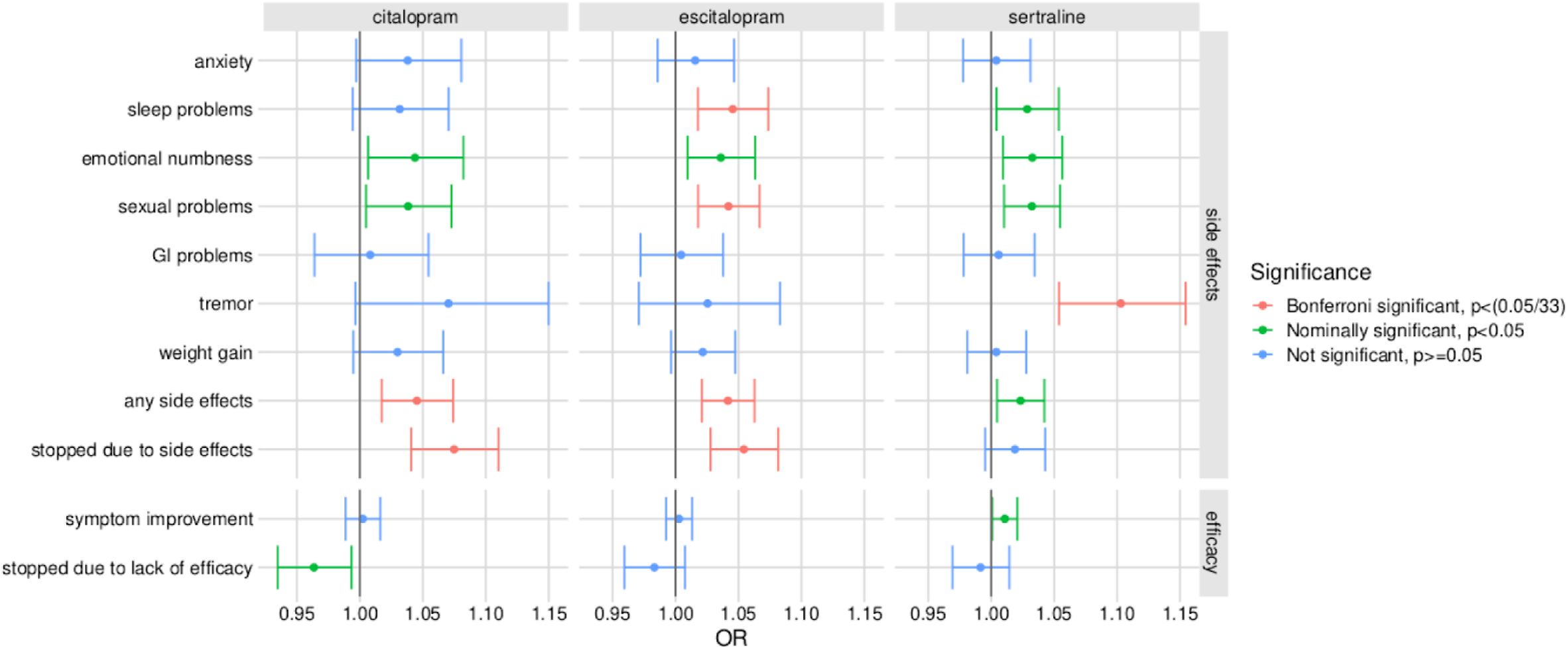
Associations between CYP2C19 metabolizer status and SSRI response. Data from cross-ancestry meta-analysis. Horizontal bars represent 95% confidence intervals. Bars are colored based on whether an association is nominally significant (green) and whether or not it survives multiple-testing correction (Bonferroni significant, red). The Bonferroni-adjusted significance threshold was conservatively defined as 0.05/k, where k represents the total number of tests conducted, 33. Symptom improvement refers to participants reporting symptoms that were either “a little” or “much” improved, as opposed to unchanged or worsened.

Associations in the European sample generally mirrored those in the meta-analyzed, cross-ancestry sample (**Supplementary Fig. 1**). We observed no Bonferroni significant associations in other genetic ancestry groups, in which we had reduced statistical power to detect associations due to smaller sample sizes (**Table 1**). The observed directions of effect were largely consistent with those in the European cohort. One notable exception was sexual problems: while faster metabolizers who used sertraline appeared less likely to report sexual problems in the European-ancestry sample (OR=1.04, 95%CI=[1.01-1.06]), they appeared more likely to do so in the African American ancestry sample (OR=0.88, 95%CI=[0.77-0.99]).

### CYP2C19 metabolizer status predicts reason for SSRI treatment discontinuation

Users of both escitalopram and citalopram who were slower CYP2C19 metabolizers were ultimately significantly more likely to discontinue treatment due to side effects (OR=1.05, 95%CI=[1.03-1.08], e.g. 29.7% vs. 21.6% and OR=1.07, 95%CI=[1.04-1.11], e.g. 25.7% vs. 20.2%, respectively, **Fig. 2**). We did not observe that faster metabolizers were significantly more likely to discontinue treatment due to a lack of efficacy in this analysis. While CYP2C19 metabolizer status clearly impacts side effect risk in our analysis, we did also not observe a significant association between CYP2C19 metabolizer status and the percentage of individuals reporting depression symptom improvement.

## Discussion

This study links genetic variation in CYP2C19 to side effects experienced by people taking SSRIs — citalopram, escitalopram and sertraline — in a large, multi-ancestry cohort. Specifically, we find that slower metabolizers of citalopram and escitalopram, as determined by their CYP2C19 genotype, are significantly more likely to experience side effects than faster metabolizers, and ultimately significantly more likely to discontinue treatment due to experiencing side effects. Owing to the large sample size of this study, we were also powered to identify association between CYP2C19 metabolizer status and specific side effects for individual drugs, where previous work typically pooled data (across drugs or side effects), and remained underpowered to detect such effects.^16,17^ We find that slower CYP2C19 metabolizers using sertraline are significantly more likely to develop tremor, as well as sexual side effects within the European-descent sample, and that slower metabolizers taking escitalopram are significantly more likely to report sleep problems and sexual problems. These genetic insights lend further support to findings from randomized clinical trials investigating pharmacogenetics-guided treatment, while circumventing the limitations of such studies (e.g., smaller scale, placebo effect).^14,15,26,27^

Analyses were limited by data availability in two important ways. First, we did not obtain drug dosage information, and were therefore unable to correct for its effect on drug response. Following standard titration, i.e., titration based on empirical side effect reporting and depression symptom control by the patient, this dosage may end up being lower for slower metabolizers (who may report more side effects) than for faster metabolizers. In other words, some of the variance in drug response that is due to genetic variation may have been reduced through titration already. In this case, we may have primarily captured the residual variation in drug response after (imperfect) titration, which would render our estimates of the effect of CYP2C19 genotype on drug response (at a constant dose) conservative.

As a second limitation, we inferred metabolizer status using only the three most common CYP2C19 alleles. Some individuals we classified as normal metabolizers may, in reality, harbor function-altering variation that we did not detect. Noisy estimates of metabolizer status could induce regression dilution, meaning our estimates of the effect of metabolizer status on treatment response may be biased downwards.

The results of this study demonstrate that self-reported drug response data are a reliable data source for genetic insights, as they align with previous observations in the pharmacogenetic literature based on other data types (e.g. rapid metabolizers of (es)citalopram having lower systemic concentrations, genetics-guided treatment reducing adverse event rates in clinical care).^26,28,29^ Self-reported data, while susceptible to selection and recall bias, are relatively simple to collect at scale and have, to date, been a powerful asset for genetic association studies; particularly for studies of disease phenotypes, where they produce findings that correlate well with those obtained from clinical (electronic health record-derived) data.^30–32^ At the time of writing, the sparsity of drug-response data in popular biobanks has precluded previous analyses of this scale. While UK Biobank, for example, has offered access to genetic data on hundreds of thousands of individuals since 2017, data collection on the efficacy of citalopram, as a popular SSRI in the UK, did not begin until late 2023, lagging behind data collection on disease phenotypes in terms of time and scale. We expect that widespread collection of self-reported drug response phenotypes in genomic cohorts will help overcome sample size limitations that have hindered genetic association studies of drug response.

Taken together, our findings demonstrate that variation in CYP2C19 predicts side effects and discontinuation due to side effects among individuals taking escitalopram, citalopram and sertraline. These findings underline the potential of pharmacogenetic testing to enable personalized therapeutic approaches that minimize side effects and treatment discontinuation – a growing need given rising rates of depression diagnoses.

## Acknowledgements

We thank the research participants and employees of 23andMe for making this work possible. The following members of the 23andMe Research Team contributed to this study: Stella Aslibekyan, Adam Auton, Elizabeth Babalola, Robert K. Bell, Jessica Bielenberg, Ninad S. Chaudhary, Zayn Cochinwala, Sayantan Das, Emily DelloRusso, Payam Dibaeinia, Sarah L. Elson, Nicholas Eriksson, Chris Eijsbouts, Teresa Filshtein, Pierre Fontanillas, Davide Foletti, Will Freyman, Zach Fuller, Julie M. Granka, Chris German, Éadaoin Harney, Alejandro Hernandez, Barry Hicks, David A. Hinds, M. Reza Jabalameli, Ethan M. Jewett, Yunxuan Jiang, Sotiris Karagounis, Lucy Kaufmann, Matt Kmiecik, Katelyn Kukar, Alan Kwong, Keng-Han Lin, Yanyu Liang, Bianca A. Llamas, Aly Khan, Steven J. Micheletti, Matthew H. McIntyre, Meghan E. Moreno, Priyanka Nandakumar, Dominique T. Nguyen, Jared O’Connell, Steve Pitts, G. David Poznik, Alexandra Reynoso, Shubham Saini, Morgan Schumacher, Leah Selcer, Anjali J. Shastri, Jingchunzi Shi, Suyash Shringarpure, Keaton Stagaman, Teague Sterling, Qiaojuan Jane Su, Joyce Y. Tung, Susana A. Tat, Vinh Tran, Xin Wang, Wei Wang, Catherine H. Weldon, Amy L. Williams, Peter Wilton. We are particularly grateful to Michael Holmes for helpful comments and suggestions.

## Data Availability

Test statistics for this study are fully disclosed in the manuscript. Individual-level data are not publicly available due to participant confidentiality, and in accordance with the IRB-approved protocol under which the study was conducted.

