## Supplementary Note for "Large-scale analysis demonstrates the influence of CYP2C19 genotype on specific SSRI side effects"

### Survey Questions about Depression Treatments

All participants who had ever taken escitalopram, citalopram or sertraline were asked about symptom improvement, side effects, and, if applicable, reasons for medication discontinuation. Participants who reported experiencing side effects were asked to select one or more side effects from a list. Participants who had used these treatments in the past were asked to select one or more reasons why they discontinued treatment from a list covering side effects, lack of symptom remission, cost, or symptom remission.

Below, we list a selection of questions used to collect data about these topics, along with preliminary questions to establish that a research participant had been diagnosed with MDD and had taken one of the three SSRIs evaluated. For clarity, instead of repeating the drug response questions for each SSRI, we show questions that pertain to citalopram. Where applicable, survey logic indicating under what circumstances a question was asked is shown between curly braces above each question.

---

**SURVEY TITLE:** Depression Treatments
**SURVEY DESCRIPTION:** Nearly 21 million American adults have experienced at least one major depressive episode in their lifetime. Genetic and non-genetic factors may influence why people respond differently to antidepressants and other treatments for depression.

Your answers to the following questions will help increase our understanding of these factors, which could improve available treatments for depression.

3. Prior survey responses show you have been diagnosed with depression. Is this correct?

7. Have you ever been hospitalized for symptoms related to depression?
( ) Yes
( ) No

9. Have you ever received any treatment for depression (such as antidepressants, counseling, shock therapy, or lifestyle changes)?

{IF ((#9 = Yes))}

12. Have you ever been prescribed or used any of the following types of treatments for **depression**? Please select all that apply.
[ ] Oral medications, such as antidepressants
[ ] Oral supplements, such as St. John's wort
[ ] Counseling or behavioral therapy

[ ] Electroconvulsive therapy (ECT or shock therapy)
[ ] Exercise or other lifestyle changes
[ ] Other, please specify: _____________

{IF ((#12 = Oral medications, such as antidepressants))}

19. Please enter the name(s) of all of the antidepressant(s) you are taking or have ever taken for **depression**.

If your medication does not appear in the list, select 'other'.
Asendin® or amoxapine
Celexa® or citalopram
Cymbalta® or duloxetine
Desyrel®, Oleptro®, Trazorel®, or trazodone
Effexor®, Effexor XR®, or venlafaxine
Elavil® or amitriptyline
Fetzima® or levomilnacipran
Lexapro®, Cipralex®, or escitalopram
Ludiomil® or maprotiline
Luvox® or fluvoxamine
Marplan® or isocarboxazid
Nardil® or phenelzine
Norpramin® or desipramine
Pamelor® or tyline
Parnate® or tranylcypromine
Paxil®, Paxil CR®, Pexeva®, or paroxetine
Pristiq®, Khedezla®, or desvenlafaxine
Prozac® or fluoxetine
Remeron® or mirtazapine
Serzone® or nefazodone
Sinequan® or doxepin
Surmontil® or trimipramine
Tofranil® or imipramine
Trintellix®, Brintellix®, or vortioxetine
Viibryd® or vilazodone
Vivactil® or protriptyline
Wellbutrin®, Wellbutrin SR®, Wellbutrin XL®, Zyban®, or bupropion
Zoloft® or sertraline
Other

{IF ((#12 = Oral medications, such as antidepressants) AND (#19 = Celexa® or citalopram))}

34. Are you **currently** taking **Celexa® (citalopram)** for depression?

( ) Yes
( ) No
{IF ((#12 = Oral medications, such as antidepressants) AND (#34 = No) AND (#19 = Celexa® or citalopram))}

36. For how long did you take **Celexa® (citalopram)**?
( ) Less than 4 months
( ) 4 months - 1 year
( ) 1 - 5 years
( ) Over 5 years

{IF ((#12 = Oral medications, such as antidepressants) AND (#34 = No) AND (#19 = Celexa® or citalopram) AND (#36 != Less than 4 months))}

38. How did your depression symptoms change after using **Celexa® (citalopram)**, compared to before using this medication?
( ) Symptoms got much worse
( ) Symptoms got a little worse
( ) Symptoms neither worsened nor improved
( ) Symptoms improved a little
( ) Symptoms improved a lot

39. Did you have any side effects while taking **Celexa® (citalopram)**?
( ) Yes
( ) No

{IF ((#12 = Oral medications, such as antidepressants) AND (#34 = No) AND (#19 = Celexa® or citalopram) AND (#39 = Yes))}

40. Which types of side effects did you have while taking **Celexa® (citalopram)**? Please select all that apply.
[ ] Anxiety
[ ] Gastrointestinal problems (such as nausea, vomiting, or dry mouth)
[ ] Numb or flattened emotions
[ ] Sexual problems
[ ] Sleep problems
[ ] Tremor
[ ] Weight gain
[ ] Other, please specify

{IF ((#12 = Oral medications, such as antidepressants) AND (#34 = No) AND (#19 = Celexa® or citalopram))}

41. Why did you stop taking **Celexa® (citalopram)**? Please select all that apply.

[ ] Side effects
[ ] My depression symptoms **did not** improve enough
[ ] Cost or insurance coverage
[ ] My depression symptoms got better

{IF ((#12 = Oral medications, such as antidepressants) AND (#34 = Yes) AND (#19 = Celexa® or citalopram))}

42. For how long have you taken **Celexa® (citalopram)**?
( ) Less than 4 months
( ) 4 months - 1 year
( ) 1 - 5 years
( ) Over 5 years

{IF ((#12 = Oral medications, such as antidepressants) AND (#34 = Yes) AND (#19 = Celexa® or citalopram) AND (#42 != Less than 4 months))}

44. How have your depression symptoms changed after using **Celexa® (citalopram)**, compared to before using this medication?
( ) Symptoms got much worse
( ) Symptoms got a little worse
( ) Symptoms neither worsened nor improved
( ) Symptoms improved a little
( ) Symptoms improved a lot

45. Have you had any side effects while taking **Celexa® (citalopram)**?
( ) Yes
( ) No
[CONCEPT: trd22_citalopram_se_curr_any_l]

{IF ((#12 = Oral medications, such as antidepressants) AND (#34 = Yes) AND (#19 = Celexa® or citalopram) AND (#45 = Yes))}

46. Which types of side effects have you had while taking **Celexa® (citalopram)**? Please select all that apply.
[ ] Anxiety
[ ] Gastrointestinal problems (such as nausea, vomiting, or dry mouth)
[ ] Numb or flattened emotions
[ ] Sexual problems
[ ] Sleep problems
[ ] Tremor
[ ] Weight gain
[ ] Other, please specify: _____________

### Supplementary Figures

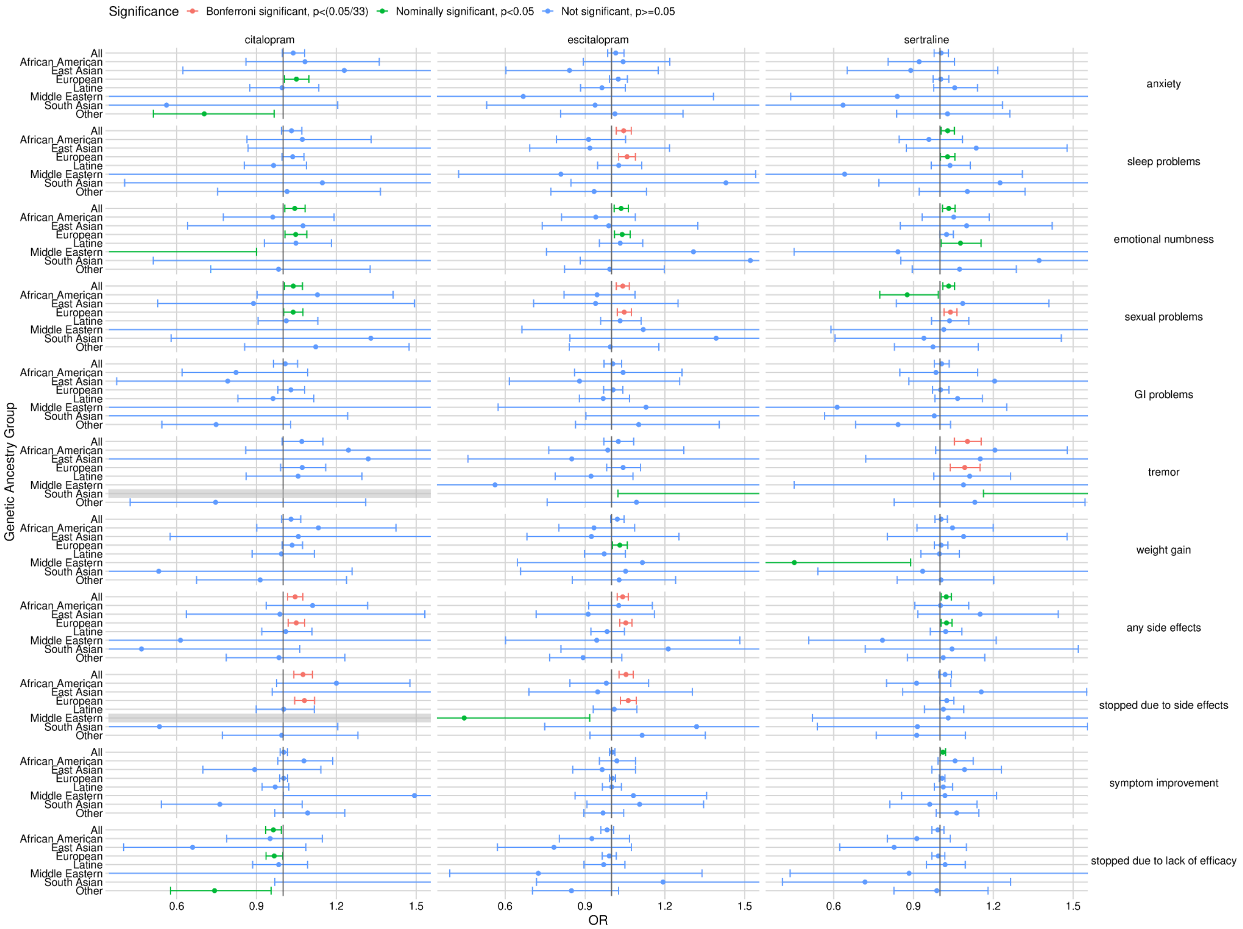

**Supplementary Figure 1: Associations between CYP2C19 metabolizer status and SSRI response by genetic ancestry group.** Horizontal bars represent 95% confidence intervals, and may extend beyond the domain shown. Grey bars indicate model fitting failures (methods). Otherwise, the color scheme and significance thresholds are identical to those in Fig. 2.
